# Nomogram for the prediction of in-hospital incidence of acute respiratory distress syndrome in patients with acute pancreatitis

**DOI:** 10.1101/2020.02.09.20019513

**Authors:** Ning Ding, Cuirong Guo, Kun Song, Changluo Li, Yang Zhou, Guifang Yang, Xiangping Chai

## Abstract

**Objective:** Acute respiratory distress syndrome (ARDS) associated with high mortality is the common complication in acute pancreatitis(AP). The aim of this study was to formulate and validate an individualized predictive nomogram for in-hospital incidence of ARDS in AP patients.

**Method:** From January 2017 to December 2018, 779 individuals with AP were included in this study. They were randomly distributed into primary cohort (n=560) and validation cohort (n=219). Based on the primary cohort, risk factors were identified by logistic regression model and a nomogram was performed. The nomogram was validated in the primary and validation cohort by the bootstrap validation method. The calibration curve was applied to evaluate the consistency between nomogram and ideal observation.

**Results:** There were 728 patients in the non-ARDS group and 51 in the ARDS group, with an incidence rate of about 6.55%. Five independent factors including white blood counts(WBC), prothrombin time(PT), albumin(ALB), serum creatinine(SCR) and triglyceride (TG) were associated with in-hospital incidence of ARDS in AP patients. A nomogram was constructed based on the five independent factors with primary cohort of AUC 0.821 and validation cohort of AUC 0.822. Calibration curve analysis indicated that the predicted probability was in accordance with the observed probability in both primary and validation cohorts.

**Conclusions:** The study developed an intuitive nomogram with easily available laboratory parameters for the prediction of in-hospital incidence of ARDS in patients with AP. The incidence of ARDS for an individual patient can be fast and conveniently evaluated by our nomogram.

## 1. Introduction

Acute pancreatitis (AP) is one of common illnesses of the gastrointestinal system, of which some can develop into severe acute pancreatitis (SAP) with poor prognosis[1]. SAP is known to be characterized by persistent organ disfunction or pancreatic necrosis, of which the mortality varies from 10% to 50%[2].

Acute respiratory distress syndrome (ARDS), as a syndrome of bilateral pulmonary infiltrates and inflammation associated with increased permeability of lung epithelium and vascular endothelium[3], occurs in around one-third of SAP patients[4]and accounts for over 50% of deaths in SAP[5]. Research evidence shows that ARDS, to a certain extent, could be preventable, and clinical prognosis may be improved due to appropriate interventions in early phase of ARDS[6]. Therefore, it is significant for the early prediction of in-hospital ARDS incidence in patients with AP.

Nomogram, as a statistical model created on the basis of different clinical and biological variables, is widely utilized for the prediction of complication, prognosis and survival in different disorders[7, 8]. And It is also beneficial for physicians to make individualized treatment plans.

However, there hasn’t been any reports on nomogram for the early prediction of ARDS in patients with AP. In this study, we analyzed the association of clinical characteristics and laboratory variables with the incidence of ARDS in AP. Furthermore, we formulated an easily applicable nomogram which included several routine laboratory parameters for predicting in-hospital incidence of ARDS in AP patients and validated its predictive capability and accuracy in an independent cohort.

## 2. Methods

### Patients

This study was a retrospect cohort study. All patients with AP admitted to the Changsha Central Hospital of University of South China and the Second Xiangya Hospital of Central South University between January 2017 and December 2018 were included. Inclusion criteria were set as follows: age ≥18 and confirmed diagnosis of AP. Excluded criteria were set as follows: more than 72 hours after onset of symptoms, recurrent pancreatitis, COPD, malignant tumors, chronic kidney disease, acute or chronic heart failure, pregnancy and HIV/ AIDS or presence of other immune-deficiency disorders.

### Ethics approval and consent to participate

The study was approved by the Ethics Committees of Changsha Central Hospital of University of South China (Changsha, China) and the Second Xiangya Hospital of Central South University (Changsha, China). All research was performed in accordance with the relevant guidelines and regulations. Due to retrospective characteristics of the study, informed consent was waived, which was also approved by the Ethics Committee of Changsha Central Hospital of University of South China (Changsha, China) and the Ethics Committee of Second Xiangya Hospital of Central South University (Changsha, China).

### Definitions

A patient was diagnosed with AP when at least two of the following criteria were encountered:1) symptom of abdominal pain, 2) the level of serum lipase or amylase increased at least three times more than the normal threshold, and 3) image findings of AP on abdominal ultrasonic and/or CT scan[2]. Hypertriglyceridemia associated with AP was defined as follows: the level of triglyceride≥ 11.3 mmol/L or ≥5.65mmol/L accompanied with milky serum[2].

Systemic inflammatory response syndrome(SIRS)was defined when two of following criteria were fulfilled: body temperature<36.0 or >38.0 °C, white blood cell count <4*10^9^/l or >12*10^9^/l or >10% immature forms, heart rate >90/min, and respiratory rate> 20/min or PaCO2< 32 mmHg[9].

According to Berlin definition of ARDS[10], the diagnosis of ARDS was made by an acute hypoxemia, a decrease in the PaO2/FiO2 index below 300 mmHg and bilateral lung infiltration on X-ray/CT scan that was not illuminated totally by fluid overload or cardiac failure. Based on PaO2/FiO2 and PEEP index, ARDS includes 3 categories: mild (200mmHg<PaO2/FiO2≤300mmHg, PEEP≥5cmH2O), moderate (100mmHg < PaO2/FiO2 ≤200 mmHg, PEEP≥5cmH2O), and severe (PaO2/FiO2≤100 mmHg, PEEP≥10cmH2O). Arterial blood gas analysis was dynamically performed for patients as well as when patients had the symptom of dyspnea during hospitalization.

### Data collection

Data collected included clinical characteristics and laboratory findings while patients were admitted in ≤24h.Collected clinical data were included : age, gender, smoking, hypertension, diabetes mellitus, coronary heart disease, etiology(alcohol, biliary, hypertriglyceridemia),systolic blood pressure(SBP),diastolic blood pressure(DBP), heart rate (HR), respiratory rate(RR), body temperature, oxyhemoglobin saturation (SaO2), white blood cell count(WBC),neutrophil(N),lymphocyte(L), hematocrit (HCT), platelet(PLT), fasting blood sugar (FBS), prothrombin time(PT), activated partial thromboplastin time(APTT), albumin (ALB), alanine aminotransferase (ALT), blood urea nitrogen(BUN),serum creatinine (SCR), amylase(AMS), lipase(LPS), lactate dehydrogenase(LDH), calcium, high density lipoprotein, (HDL), low density lipoprotein(LDL), triglyceride(TG), PaO2 and PaCO2. NLR is defined as the ratio of neutrophils to lymphocytes. Incidence of SIRS within 24 hours after admission was calculated. The scores of sequential organ failure assessment (SOFA), bedside index of severity in acute pancreatitis (BISAP),and Ranson were calculated to assess the severity of AP patients in terms of personal characteristics, vital signs and laboratory findings (Table 1).

**Table 1.** General characteristics of 779 patients with AP.

|  | non-ARDS(n=728) | ARDS(n=51) | P-value |
| --- | --- | --- | --- |
| **Demographics** |  |  |  |
| Male, n (%) | 509(69.92%) | 42(82.35%) | 0.059 |
| Age, median( IQR)(years) | 45(36-55) | 41(35-51) | 0.617 |
| Diabetes mellitus, n(%) | 111(15.25%) | 8(15.69%) | 0.933 |
| Hypertension, n(%) | 134(18.41%) | 9(17.65%) | 0.892 |
| Coronary heart disease, n(%) | 23(3.16%) | 2(3.92%) | 0.766 |
| Smoking, n(%) | 262(35.99%) | 23(45.1%) | 0.067 |
| **Etiology** |  |  |  |
| Alcohol, n(%) | 137(18.82%) | 12(23.53%) | 0.409 |
| Hypertriglyceridemia, n(%) | 283(38.87%) | 29(56.86%) | 0.011 |
| Biliary, n(%) | 148(20.33%) | 6(11.76%) | 0.138 |
| **Vital signs** |  |  |  |
| SBP, median (IQR)(mmHg) | 130(121-142) | 132(120-144) | 0.342 |
| DBP, median (IQR)(mmHg) | 81(75-89.75) | 82(75-90) | 0.919 |
| HR, median(IQR)(beats/min) | 80(74-88) | 99(80-116) | <0.001 |
| RR, median (IQR)(beats/min) | 18(17-19) | 18(17-20) | 0.125 |
| SaO<sub>2</sub>, median(IQR)(%) | 96(96-97) | 98(94-98) | 0.225 |
| Temperature (°C ) | 36.7(36.5-36.9) | 36.9(36.8-37.3) | 0.025 |
| **Clinical outcomes** |  |  |  |
| Ventilation, n(%) | 0(0%) | 19(37.25%) | <0.001 |
| In-hospital mortality, n(%) | 0(0%) | 3(5.882%) | <0.001 |
| LOS in hospital ,<br>median(IQR)(days) | 7(5-9) | 12(8-19) | <0.001 |
| LOS in ICU,<br>median (IQR)(days) | 0(0-0) | 6(4-8) | <0.001 |
| **Laboratory findings** |  |  |  |
| WBC, median (IQR)(<sup>^</sup>10<sup>9</sup>/l) | 12.32(9.27-15.15) | 15.425(12.36-18.20) | <0.001 |
| N, median (IQR)(<sup>^</sup>10<sup>9</sup>/l) | 9.76(6.65-12.51) | 12.46(9-14.84) | <0.001 |
| L, median (IQR)(<sup>^</sup>10<sup>9</sup>/l) | 1.49(1.01-2.16) | 1.19(0.8-2.02) | 0.025 |
| NLR, median (IQR) | 6.25(3.61-10.41) | 11.24(6.20-19.92) | 0.001 |
| HCT, median (IQR) | 0.43(0.40-0.47) | 0.46(0.41-0.48) | 0.006 |
| PLT, median (IQR)(<sup>^</sup>10<sup>9</sup>/l) | 209(167-250) | 211(174-253.25) | 0.808 |
| FBS, median (IQR)(mmol/l) | 7.4(6.4-9.5) | 9.8(7.77-13.45) | <0.001 |
| PT, median (IQR)(s) | 11.4(10.7-12.2) | 12.75(11.7-14.42) | <0.001 |
| APTT, median (IQR)(s) | 27.8(24.3-31.75) | 32.1(26.52-36.12) | <0.001 |
| ALB, median (IQR)(g/l) | 43.7(40.2-46.9) | 39.9(36.2-46.6) | 0.049 |
| ALT, median (IQR)(u/l) | 33(20-71.12) | 40(22.9-87) | 0.683 |
| BUN, median (IQR)(mmol/l) | 4.7(3.67-5.99) | 4.9(4.33-6.61) | 0.017 |
| SCR, median (IQR)(umol/l) | 66.95(54.22-77) | 76(59-123) | 0.012 |
| AMS, median (IQR)(u/l) | 310.5(113.25-954.75) | 369(114-1150) | 0.932 |
| LPS, median (IQR)(u/l) | 385(124-859.5) | 404(221-876) | 0.570 |
| LDH, median (IQR)(u/l) | 228(180-304.75) | 398(219-543) | <0.001 |
| Calcium, median<br>(IQR)(mmol/l) | 2.4(2.3-2.5) | 2.34(2.07-2.51) | 0.024 |
| HDL, median (IQR)(mmol/l) | 0.92(0.65-1.3) | 0.61(0.46-0.91) | 0.089 |
| LDL ,median (IQR)(mmol/l) | 2.18(1.39-2.96) | 1.48(1.09-2.438) | 0.928 |
| TG, median (IQR)(mmol/l) | 5.59(1.43-17.77) | 17.16(2.56-26.69) | 0.003 |
| PaO<sub>2</sub>, median (IQR)(mmHg) | 78.5(75.3-82.5) | 77.1(74.6-81.8)- | 0.068 |
| PaCO<sub>2</sub>, median (IQR)(mmHg) | 38.8(36-41.5) | 37.4(36.3-41.6) | 0.085 |
| SIRS, n(%) | 276(37.9%) | 26(50.98%) | 0.026 |
| SOFA | 0(0-1) | 2(1-3) | <0.001 |
| RANSON | 1(0-2) | 4(2-5) | <0.001 |
| BISAP | 1(0-2) | 2(2-3) | <0.001 |

**Abbreviations:** ICU, Intensive care unit; LOS, length of stay; SBP, systolic blood pressure; DBP, diastolic blood pressure; HR, heart rate; RR, respiratory rate ,SaO<sub>2</sub>, oxyhemoglobin saturation; WBC, white blood cell count; N, neutrophil; L, lymphocyte; HCT, hematocrit; PLT, platelet; FBS, fasting blood sugar; PT, prothrombin time; APTT, activated partial thromboplastin time; ALB, albumin; ALT, alanine aminotransferase; BUN, blood urea nitrogen; SCR, serum creatinine; AMS, amylase; LPS, lipase; LDH, lactate dehydrogenase; HDL, high density lipoprotein; LDL, low
density lipoprotein; TG, triglyceride; SOFA, Sequential organ failure assessment; BISAP, bedside index of severity in acute pancreatitis; SIRS, systemic inflammatory response syndrome; NLR is defined as the ratio of neutrophils to lymphocytes.

Primary clinical outcome was in-hospital incidence of ARDS. Length of stay(LOS) in ICU and hospital were also compared. (Table 1).

### Statistical analysis

Statistical results were expressed in mean ± standard deviation for normal data, while for non-normal data, interquartile range (IQR)and median were employed. Categorical data were expressed as percentage and number. Student t-test or Rank sum test was applied for comparisons between ARDS and non-ARDS groups. Chi-square or Fishers exact test was applied for categorical data.

The original database was randomly distributed into the primary and validation cohort by 7:3 ratio. The primary cohort was applied to develop the model by multivariable logistic regression and the validation cohort was applied to validate the model. The area under receiver operating characteristic curve (AUC) was utilized for evaluating the accuracy of the model.

A nomogram was performed on the basis of logistic regression model and we proportionally converted each regression coefficient in the model to a scale of 0 to 100point. The total points were calculated by adding the point of each independent variable, which predicted the individualized incidence of ARDS. The nomogram was validated in the primary and validation cohort by the bootstrap validation method. The calibration curve was applied to evaluate the consistency between nomogram and observation probability.

Statistical analysis was performed using SPSS software (version 26) and R software (http://www.R-project.org). Statistical significance was considered to be at two-sided p < 0.05.

## 3 Results

### General Characteristics of all patients

A total of 817 patients with diagnosis of AP were initially identified and 38 were excluded according to the excluded criteria. Finally, a total of 779 patients were included to analyze in the study.(Figure 1)

**Figure 1.**
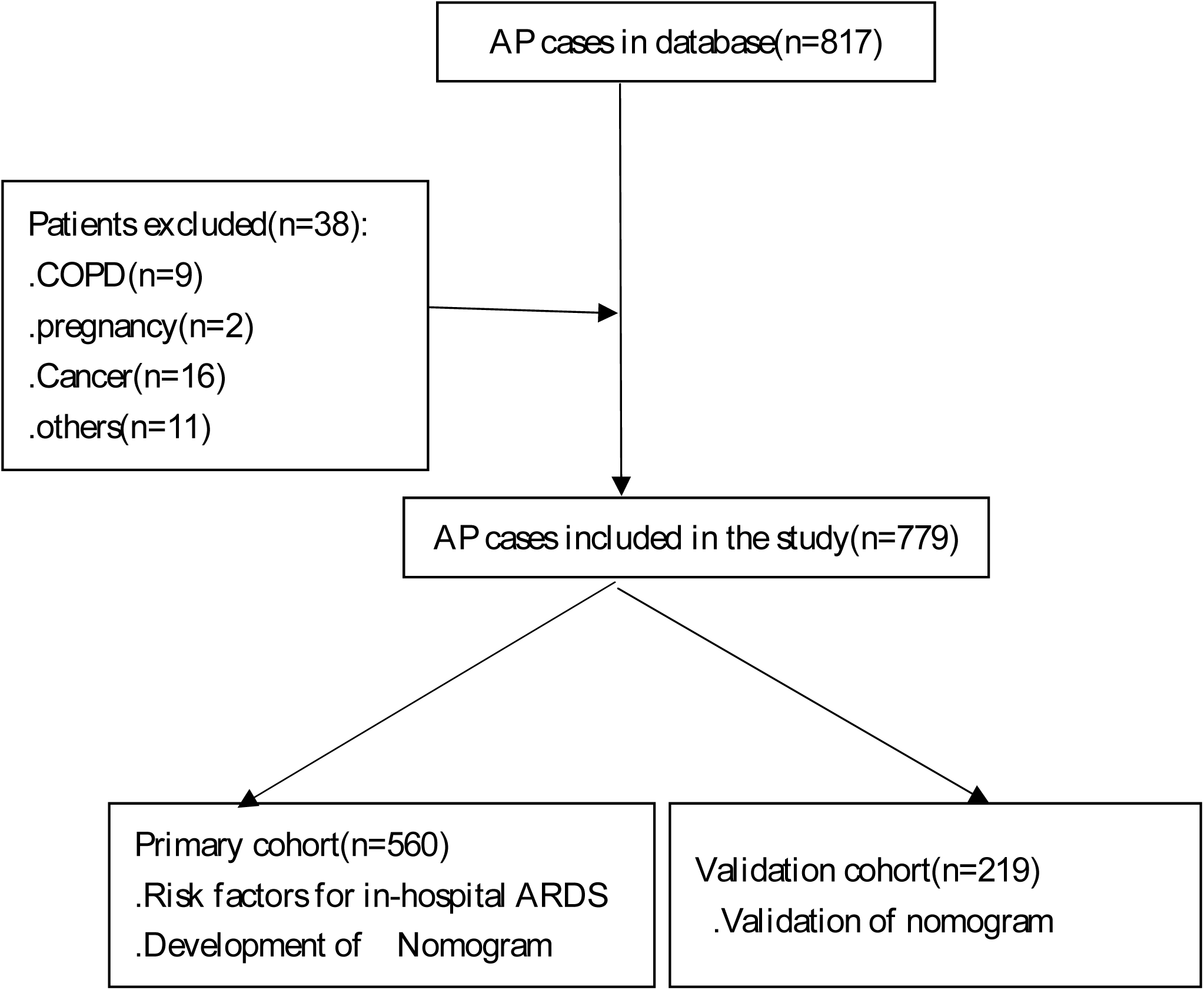
Flow diagram of patients enrollment and study design.

The general characteristics of the patients were showed in Table 1. There were 728 patients in the non-ARDS group and 51 in the ARDS group, with an incidence rate of about 6.55%. In ARDS group, the number of mild, moderate and severe ARDS were 32, 16 and 3, respectively. Proportion of males in ARDS group was 82.35% while 69.92% in non-ARDS group. Age at the onset, smoking, past medical history (diabetes mellitus, hypertension, coronary heart disease), etiology(alcohol, biliary), SBP, DBP, RR and SaO2 were not significantly different between patients with and without ARDS. Hypertriglyceridemia(P=0.011), temperature(P=0.025) and heart rate(P<0.001) were significantly different between two groups (both p < 0.05).

For ARDS group, ALB was significant lower than that in non-ARDS group(P=0.049). Other laboratory tests such as WBC(P<0.001),N(P<0.001),L(P=0.025), NLR(P=0.001), HCT(P=0.006),FBS(P<0.001),PT(P<0.001),APTT(P<0.001),BUN(P=0.017),SCR (P=0.012), LDH(P <0.001),calcium(P=0.024) and TG(P=0.003) were significantly higher in the ARDS group than that in the non-ARDS group. PLT, ALT, AMS, LPS, HDL, LDL, PaO2 and PaCO2 were not significantly different between two groups. The incidence of SIRS within 24 hours after admission was significant higher in ARDS group(P=0.026).(Table 1)

The incidence of ventilation and in-hospital mortality in ARDS group were 37.25%(19/51) and 5.882%(3/51), respectively. LOS in ICU and hospital (all P <0.001) were also significantly higher in the ARDS group. Scores of SOFA, Ranson and BISAP (all P<0.001) were significantly higher in the ARDS group than that in the non-ARDS group. (Table 1)

### Baseline characteristics of primary and validation cohort

The baseline characteristics of patients in the primary cohort(n=560) and validation cohort(n=219) are shown in Table 2. Proportion of males in primary and validation cohorts were respectively 68.57% and 76.25%(P=0.378). The main etiologies of AP in primary cohort were hypertriglyceridemia(40.17%), alcohol(19.64%) and biliary (18.21%),while in validation cohort the etiologies were hypertriglyceridemia(39.72%), biliary(23.74%) and alcohol(17.8%). The incidence of ARDS were respectively 6.446% and 6.829% in two groups(P=0.416).

**Table 2.** Comparison primary cohort and validation cohort.

|  | Primary cohort<br>(n=560) | Validation cohort<br>(n=219) | P-value |
| --- | --- | --- | --- |
| **Demographics** |  |  |  |
| Male, n (%) | 384(68.57%) | 167(76.25%) | 0.378 |
| Age, median( IQR)(years) | 45(36-55) | 45(36-57) | 0.094 |
| Diabetes, n (%) | 89(15.89%) | 30(13.69%) | 0.001 |
| Hypertension, n (%) | 106(18.92%) | 37(16.89%) | 0.133 |
| Coronary heart disease, n(%) | 22(3.92%) | 3(1.36%) | 0.04 |
| Smoking n(%) | 199(35.53%) | 86(39.27%) | 0.402 |
| **Etiology** |  |  |  |
| Alcohol, n (%) | 110(19.64%) | 39(17.80%) | 0.02 |
| Hypertriglyceridemia, n (%) | 225(40.17%) | 87(39.72%) | 0.081 |
| Biliary, n (%) | 102(18.21%) | 52(23.74%) | 0.148 |
| **Vital signs** |  |  |  |
| SBP, median (IQR)(mmHg) | 130(120-142) | 132(122-142) | 0.76 |
| DBP, median (IQR)(mmHg) | 81(74-90) | 82(75-89) | 0.258 |
| HR, median (IQR)(beats/min) | 80(74-90) | 80(75-90) | 0.418 |
| RR, median (IQR)(beats/min) | 18(17-19) | 18(17-20) | 0.422 |
| SaO<sub>2</sub>,median(IQR)(%) | 96(96-97) | 97(96-97) | 0.356 |
| Temperature(°C) | 36.7(36.5-36.9) | 36.7(36.6-37) | 0.525 |
| ARDS, n (%) | 36(6.446%) | 15(6.829%) | 0.416 |
| **Laboratory findings** |  |  |  |
| WBC, median (IQR)(<sup>^</sup>10<sup>9</sup>/l) | 12.49(9.42-15.4) | 12.55(9.43-15.52) | 0.13 |
| N, median (IQR)(<sup>^</sup>10<sup>9</sup>/l) | 9.91(6.82-12.76) | 10.16(6.83-12.58) | 0.517 |
| L, median (IQR)(<sup>^</sup>10<sup>9</sup>/l) | 1.44(0.98-2.1) | 1.50(1.04-2.27) | 0.445 |
| NLR, median (IQR) | 6.74(3.683-11.415) | 6.21(3.729-9.626) | 0.606 |
| HCT, median (IQR) | 0.43(0.4-0.46) | 0.44(0.41-0.48) | 0.073 |
| PLT, median (IQR)(<sup>^</sup>10<sup>9</sup>/l) | 208(167-248) | 212(170.25-254.75) | 0.222 |
| FBS, median (IQR)(mmol/l) | 7.5(6.4-9.7) | 7.7(6.4-10) | 0.094 |
| PT, median (IQR)(s) | 11.5(10.8-12.3) | 11.4(10.7-12.3) | 0.268 |
| APTT, median (IQR)(s) | 28.1(25.2-32.5) | 27.15(23.72-31.2) | 0.147 |
| ALB, median (IQR)(g/l) | 43.5(40-46.8) | 43.7(40-46.9) | 0.072 |
| ALT, median (IQR)(u/l) | 33(20-68) | 38(21.15-81.25) | 0.986 |
| BUN, median (IQR)(mmol/l) | 4.7(3.7-5.99) | 4.905(3.76-6.42) | 0.175 |
| SCR, median (IQR)(umol/l) | 66(54-57) | 69(57.85-79.8) | 0.06 |
| AMS, median (IQR)(u/l) | 323(113.5-946) | 284(116.25-1200.35) | 0.001 |
| LPS, median (IQR)(u/l) | 398.5(132.75-862.25) | 369(126.75-868.25) | 0.004 |
| LDH, median (IQR)(u/l) | 230(180.75-313) | 233(184.5-328) | 0.217 |
| Calcium, median (IQR)(mmol/l) | 2.39(2.29-2.49) | 2.41(2.29-2.51) | 0.827 |
| HDL, median (IQR)(mmol/l) | 0.91(0.63-1.28) | 0.905(0.58-1.29) | 0.193 |
| LDL, median (IQR)(mmol/l) | 2.15(1.33-2.9) | 2.115(1.40-3.12) | 0.191 |
| TG, median (IQR)(mmol/l) | 5.88(1.43-18.6) | 5.635(1.53-21.00) | 0.699 |
| PaO<sub>2</sub>, median (IQR)(mmHg) | 78.4(75.5-81.6) | 77.9(75.2-82.1) | 0.185 |
| PaCO<sub>2</sub>, median (IQR)(mmHg) | 38.5(36.5-41.4) | 38.2(36.8-41.5) | 0.108 |
| SIRS,n(%) | 210(37.5%) | 92(42.01%) | 0.134 |

**Abbreviations:** SBP, systolic blood pressure; DBP, diastolic blood pressure; HR, heart rate; RR, respiratory rate,SaO<sub>2</sub>, oxyhemoglobin saturation; WBC, white blood cell count; N, neutrophil; L,

Except serum amylase and lipase, there were no significant difference between the primary and validation cohort in other laboratory tests.

### Comparison of variables between ARDS and non-ARDS groups in primary cohort

Variables including HR(P<0.001), temperature(P=0.027), WBC(P<0.001), N(P<0.001), L(P=0.018), NLR(P=0.004), FBS(P<0.001), PT(P<0.001), APTT(P=0.01), ALB (P<0.001), SCR(P<0.001),LDH(P<0.001), Calcium(P=0.003), HDL(P=0.034), TG (P<0.001) and SIRS(P=0.045) were significantly different between two groups in primary cohort(Table 3).

**Table 3.** Comparison of variables between ARDS and non-ARDS groups in primary cohort.

|  | non-ARDS(n=524) | ARDS(n=36) | P-value |
| --- | --- | --- | --- |
| **Demographics** |  |  |  |
| Male, n (%) | 355(67.75%) | 20(85.55%) | 0.11 |
| Age, median ( IQR)(years) | 46(36-55)- | 43(34-50.75) | 0.375 |
| Diabetes mellitus, n(%) | 84(16.03%) | 5(13.88%) | 0.726 |
| Hypertension, n(%) | 99(18.89%) | 7(19.44%) | 0.97 |
| Coronary heart disease, n(%) | 20(3.81%) | 2(5.55%) | 0.663 |
| Smoking, n(%) | 185(35.3%) | 14(38.89%) | 0.368 |
| **Etiology** |  |  |  |
| Alcohol, n(%) | 102(19.46%) | 8(22.22%) | 0.705 |
| Hypertriglyceridemia, n(%) | 206(39.31%) | 19(52.77%) | 0.13 |
| Biliary, n(%) | 98(18.7%) | 4(11.11%) | 0.254 |
| **Vital signs** |  |  |  |
| SBP, median (IQR)(mmHg) | 130(120-142) | 132(116.5-145) | 0.448 |
| DBP, median (IQR)(mmHg) | 81(74-90) | 80.5(75.25-90) | 0.964 |
| HR, median(IQR)(beats/min) | 80(74-88) | 99.5(80.25-119.5) | <0.001 |
| RR, median (IQR)(beats/min) | 18(17-19) | 19(19-20) | 0.224 |
| SaO<sub>2</sub>,median(IQR)(%) | 97(96-97) | 96(94-98) | 0.306 |
| Temperature (°C ) | 36.7(36.5-36.9) | 36.9(36.8-37.3) | 0.027 |
| **Laboratory findings** |  |  |  |
| WBC, median (IQR)(<sup>^</sup>10<sup>9</sup>/l) | 12.28(9.35-15.14) | 15.55(12.663-19.37) | <0.001 |
| N, median (IQR)(<sup>^</sup>10<sup>9</sup>/l) | 9.68(6.675-12.525) | 13.01(9.23-17.84) | <0.001 |
| L, median (IQR)(<sup>^</sup>10<sup>9</sup>/l) | 1.48(0.99-2.11) | 1.12(0.81-1.67) | 0.018 |
| NLR, median (IQR) | 6.31(3.61-10.81) | 12.34(6.78-19.33) | 0.004 |
| HCT, median (IQR) | 0.43(0.4-0.46) | 0.45(0.41-0.48) | 0.083 |
| PLT, median (IQR)(<sup>^</sup>10<sup>9</sup>/l) | 208(167-247.5) | 212(171-253.75) | 0.949 |
| FBS, median (IQR)(mmol/l) | 7.4(6.4-9.42) | 10(7.72-13.55) | <0.001 |
| PT, median (IQR)(s) | 11.4(10.8-12.2) | 12.7(11.7-14.8) | <0.001 |
| APTT, median (IQR)(s) | 28(25.02-32.1) | 32.3(26.7-36.1) | 0.01 |
| ALB, median (IQR)(g/l) | 43.7(40.3-46.8) | 39.6(34.6-46.45) | <0.001 |
| ALT, median (IQR)(u/l) | 32.7(20-64) | 40(21-129.5) | 0.297 |
| BUN, median (IQR)(mmol/l) | 4.69(3.67-5.96) | 4.82(4.34-6.85) | 0.103 |
| SCR, median (IQR)(umol/l) | 65.1(53.8-75) | 78.5(59.5-121) | <0.001 |
| AMS, median (IQR)(u/l) | 318(113-899) | 438.5(118-1219) | 0.752 |
| LPS, median (IQR)(u/l) | 397(126-843.75) | 442.5(200.25-920.5) | 0.31 |
| LDH, median (IQR)(u/l) | 226(178.75-302.25) | 403(222.5-551.25) | <0.001 |
| Calcium, median (IQR)(mmol/l) | 2.39(2.29-2.49) | 2.31(2.14-2.48) | 0.003 |
| HDL, median (IQR)(mmol/l) | 0.92(0.65-1.29) | 0.67(0.43-0.96) | 0.034 |
| LDL ,median (IQR)(mmol/l) | 2.18(1.36-2.91) | 1.515(1.128-2.33) | 0.431 |
| TG, median (IQR)(mmol/l) | 5.74(1.41-17.09) | 13.21(2.48-25.06) | <0.001 |
| PaO2, median (IQR)(mmHg) | 78.1(75-82.6) | 76.1(73.6-80.5)- | 0.087 |
| PaCO2,median (IQR)(mmHg) | 38.5(36-41.2) | 37.2(36.1-40.6) | 0.132 |
| SIRS,n(%) | 193(36.83%) | 17(47.22%) | 0.045 |

**Abbreviations:** SBP, systolic blood pressure; DBP, diastolic blood pressure; HR, heart rate; RR, respiratory rate; SaO<sub>2</sub>, oxyhemoglobin saturation; WBC, white blood cell count; N, neutrophil; L, lymphocyte; HCT, hematocrit; PLT, platelet; FBS, fasting blood sugar; PT, prothrombin time; APTT, activated partial thromboplastin time; ALB, albumin; ALT, alanine aminotransferase; BUN, blood urea nitrogen; SCR, serum creatinine; AMS, amylase; LPS, lipase; LDH, lactate dehydrogenase; HDL, high density lipoprotein; LDL, low density lipoprotein; TG, triglyceride; SOFA, Sequential organ failure assessment; BISAP, bedside index of severity in acute pancreatitis; SIRS, systemic inflammatory response syndrome; NLR is defined as the ratio of neutrophils to lymphocytes.

### Multivariate logistic regression

The model with five independent variables were identified by multivariate logistic regression analysis in primary cohort (Table 4): WBC (Odds Ratio(OR)=1.115,95%CI : 1.031-1.206,P=0.0064),PT(OR=1.412,95%CI:1.147-1.74,P=0.0011),ALB(OR= 0.925, 95%CI:0.863-0.993,P=0.0302),SCR(OR=1.007,95%CI:1.001-1.015,P=0.0348)and TG (OR=1.086, 95%CI:1.045-1.129, P<0.001).

**Table 4.**
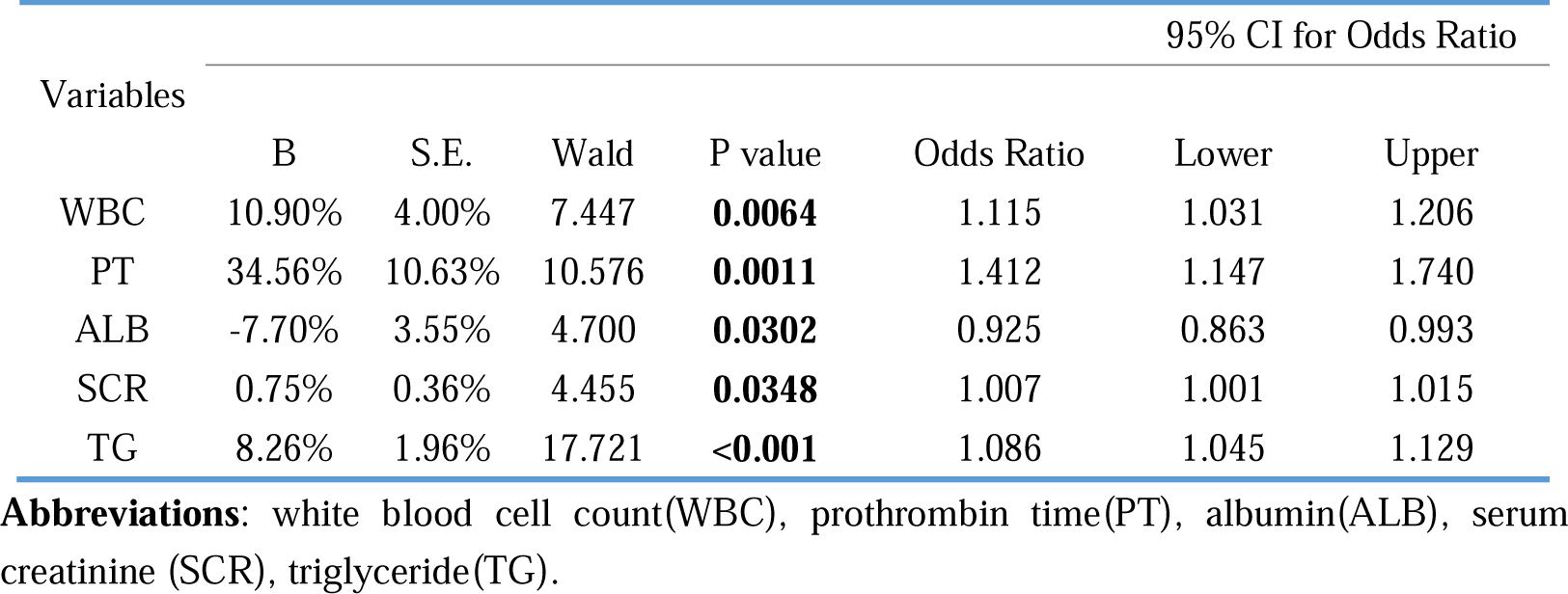
Multivariate logistic regression analysis of primary cohort.

### Nomogram

Nomogram was performed on the basis of the significantly independent and predictive variables identified by multivariate logistic regression. WBC, PT, ALB, SCR and TG were included in the predictive nomogram, which was a graphical version of the statistical model that illuminated the link between the predictive variables and the probability of in-hospital incidence of ARDS (Figure 2). The higher the score calculated in the nomogram, the higher the likelihood of incidence of ARDS. For example, clinical data of two patients in our study were analyzed. First patient with PT of 12.8(32points), WBC of 38.48(32points), TG of 13.6 (10points), ALB of 46(30points) and SCR of 166 (8 points) scored a total of 112 points. Therefore, the patient had approximate over 50% probability of developing into ARDS in hospital. Actually, he was transferred to ICU due to ARDS. Another patient with PT of 11.1(27points), WBC of 16.65(13points), TG of 26.95(15points), ALB of 47.6 (28points)and SCR of 53(3 points) scored a total of 86 points. The in-hospital incidence of ARDS was less than 0.1 and the patient recovered totally in less than one week without ARDS.

**Figure 2.**
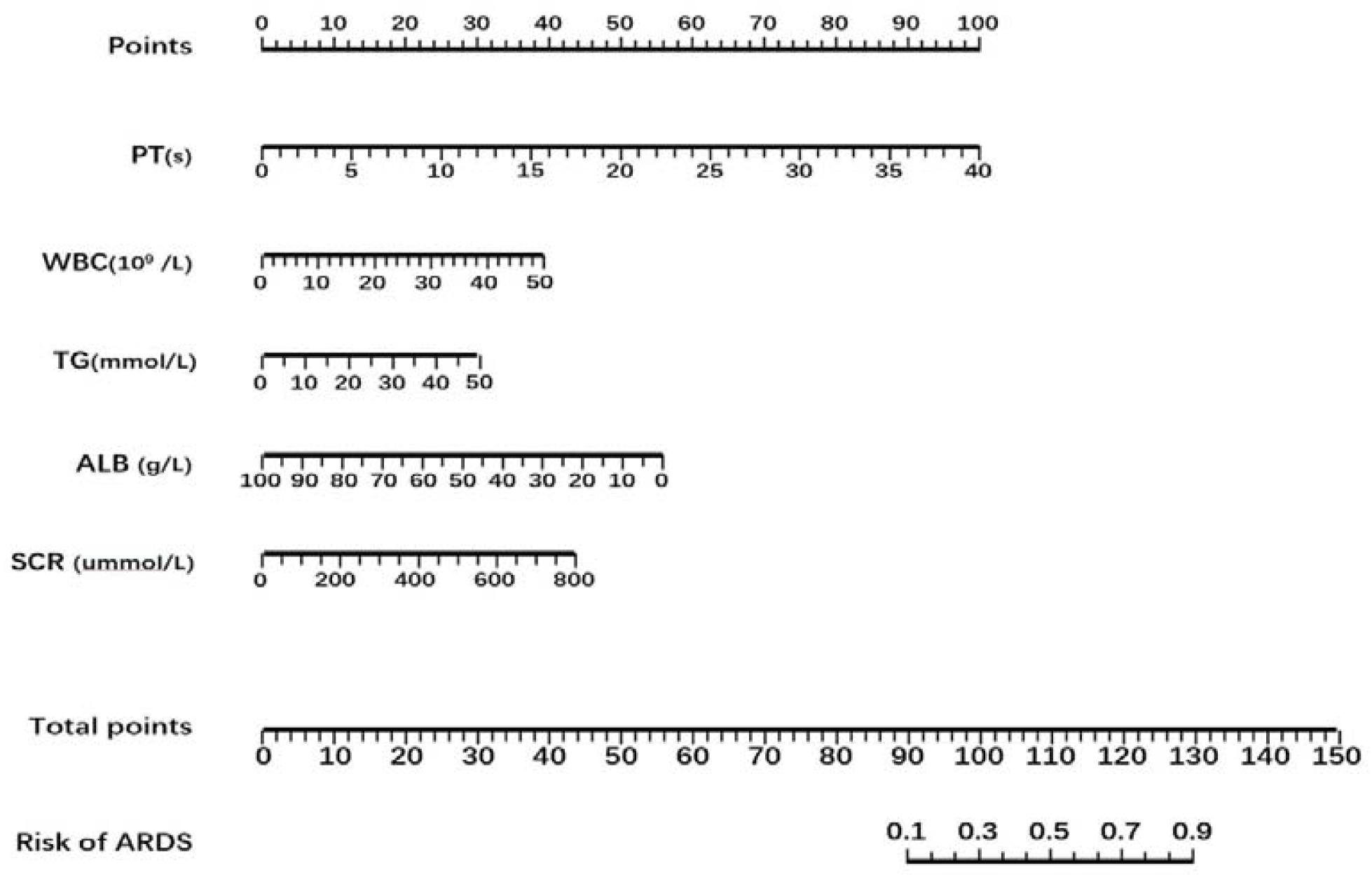
Nomogram indicating the risk of ARDS in AP. **Abbreviations**: white blood cell count(WBC),prothrombin time(PT), albumin(ALB), serum creatinine(SCR), triglyceride(TG).

### Validation of the predictive accuracy of nomogram in primary and validation cohort

In primary cohort, the nomogram displayed a good accuracy in evaluating the risk of ARDS in AP patients with an AUC=0.821(95%CI:0.756-0.886) (Figure 3A), which was more than 0.8 demonstrating a good discrimination[11]. The calibration curve showed that the predicted probability was in accordance with the observed probability(Figure 3B).

**Figure 3.**
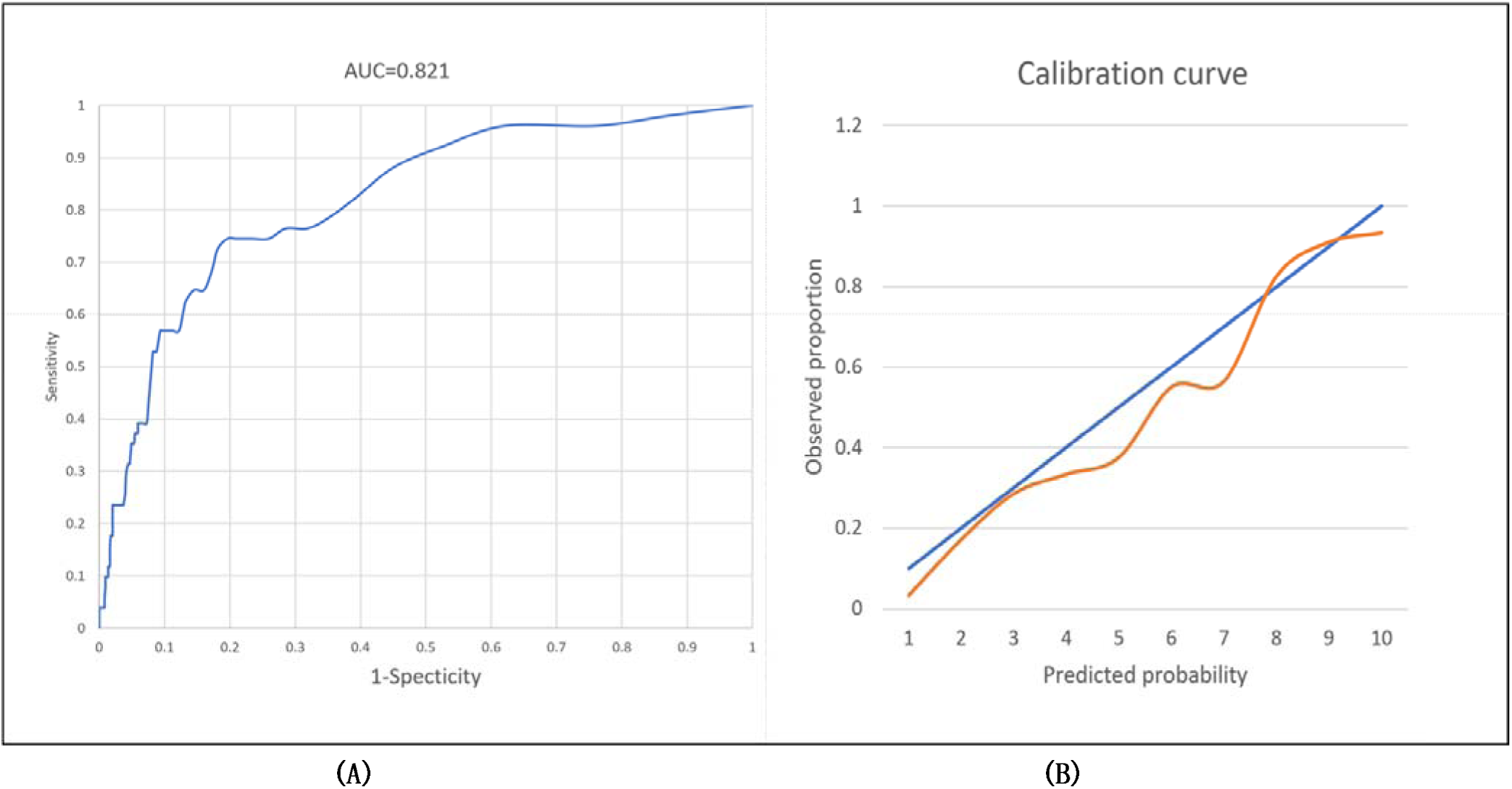
Validation of nomogram in primary cohort. (A) Discrimination: AUC of the ROC curve was 0.821 (95%CI: 0.756-0.886). (B) Nomogram calibration curve. The blue line indicates perfect prediction by an ideal model. The orange line indicates actual performance of the model.

In validation cohort, the nomogram also verified its accuracy with an AUC=0.823 (95%CI:0.707-0.937)(Figure 4A). The calibration curve also demonstrated that there was relatively close consistency between the predicted probability and the observed probability(Figure 4B).

**Figure 4.**
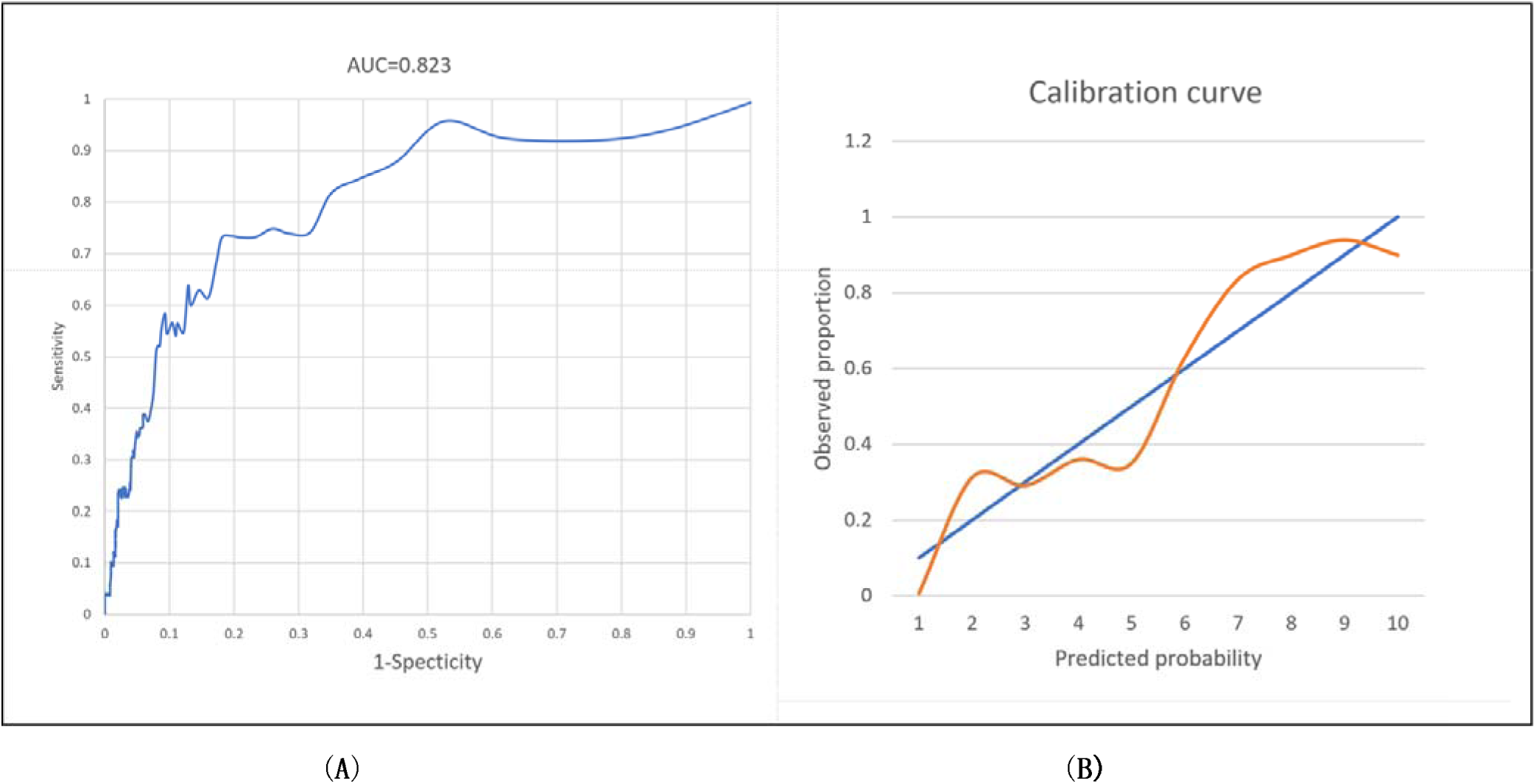
Validation of nomogram in validation cohort. (A) Discrimination: AUC of the ROC curve was 0.823 (95%CI: 0.707-0.937). (B) Nomogram calibration curve. The blue line indicates perfect prediction by an ideal model. The orange line indicates actual performance of the model.

## 4 Discussion

AP patients varied in severity of disease, clinical course and outcomes. 20-30% of AP developed into severe complications such as acute kidney injury and ARDS. ARDS associated with high mortality was the common complication in AP[12]. Early identification of ARDS in AP patients may contribute to identifying the patients who need to be monitored and ameliorating the clinical outcomes[13].

The aim of this study was to formulate and validate an individualized predictive nomogram for in-hospital incidence of ARDS in AP patients. In the study, we analyzed and evaluated the predictive capability of clinical and laboratory parameters by logistic regression. We found several different predictors incorporating WBC, PT, SCR, ALB and TG that were independently associated with the increased risk of ARDS. Therefore, predictive nomogram was constructed.

In our study, WBC counts and neutrophil counts in AP patients with ARDS were significantly higher than those without ARDS. Evidence showed that WBC as a biomarker associated with systemic inflammatory response could be potential predictors in various disorders such as cerebral vascular disease, cancer and pulmonary dysplasia[14-16]. In both gallstone AP and hypertriglyceridemia□ induced AP, WBC was also an effective indicator for predicting severity and prognosis, which was consistent with our study[17]. This phenomenon can be partly explained by that the etiology including gallstone and hypertriglyceridemia accounted for over 50% AP patients in our study. Due to the increased WBC and neutrophils in circulation, their toxic mediators can lead to lung tissue injury with an increased permeability of lung epithelium and endothelium, which finally results in respiratory failure such as ARDS[18].

The dynamic changes of coagulation such as prolonged PT and APTT were identified risk factors and related to the worse outcome of AP[19]^18^. Several studies have illuminated that systemic inflammation reaction and coagulative dysfunction played a significant role in the development of AP, which may lead to pancreatic necrosis, acute kidney injury and ARDS[20, 21].

Elevated SCR as an indicator of acute kidney injury (AKI) was associated with pancreatic necrosis, organ failure and mortality in AP as well[22, 23].Early change especially in the first 24 hours after admission in SCR level was an effective predictor of disease severity in AP[24]. In the early phase of AP, inflammatory mediators and hypoperfusion in kidney contributed to the progress of AKI, which on the contrary also increased inflammatory cytokines such as IL-6 and aggravated lung injury[25].

In the ARDS group of our study, levels of serum albumin were significantly lower than those without ARDS. Hypoalbuminemia has been found to be linked with the morbidity and mortality in many different disorders such as sepsis and AKI[26, 27].In AP patients, partly due to decreased enteral energy intake and increased tissue protein catabolism, low serum albumin was an independent indicator of worse clinical outcome[28, 29]. The lower the level of serum albumin, the higher the risk of persistent organ failure[30]. Albumin as an osmotic pressure maintainer may improve hemodynamic stabilization and decrease alveolar-capillary permeability and inflammatory reaction which can ameliorate oxygenation in patients with ARDS[31].

Hypertriglyceridemia is one of the risky factors of AP as well as in cerebral cardiovascular disease and all-cause mortality[32, 33]. Some previous reports revealed that the incidence of hypertriglyceridemia□induced AP varied from 10-30% in different countries[34, 35]. In China, the incidence of hypertriglyceridemia associated AP has been increased in recent years[36]. In our study, hypertriglyceridemia accounted for around 1/3 in the etiologies of AP. Evidence showed that an increased TG level was related to the severity and clinical outcomes of AP, including pancreatic necrosis, organ failure, ICU admission, and mortality[37]. Research illuminated that TG of excess amounts in circulation were hydrolyzed into high levels of free fatty acids(FFA) by pancreatic lipase[38]. FFA can impair vascular endothelium in microcirculation and result in an increment in viscosity which may cause hypoperfusion, inflammation, tissue ischemia, and organ dysfunction eventually[39].

As far as we know, this is the first study to develop a nomogram for predicting the in-hospital incidence of ARDS in AP patients. For each AP patient, our nomogram enables physicians to directly and conveniently calculate a numeric probability of ARDS. It also helps physicians for clearly communicating with a patient via individual nomogram evaluation and making individualized treatment plans.

In our study, there are still several limitations. First, hospitals in this study were tertiary hospitals in urban city, which may lead to the difference of clinical characteristics in AP compared to the patients in hospitals which located in the suburb of city and countryside or other areas. It could partly explained that why the etiologies and variables associated with ARDS in AP in our study differed from some other researches[40]. When applying the proposed nomogram to a larger population or other centers, caution must be used. Multicenter research with larger samples needs to be explored for validating our model. Second, the different models in three subgroups of ARDS couldn’t be performed in this study due to its relatively small samples. For better applying the model in different patient, studies should be carried out by expanding the sample size in the future. Third, our study was retrospective and there might be patient selection biases, which was an inevitable limitation in these types of studies. Moreover, due to its retrospective nature and part of data missing(such as C-reactive protein, procalcitonin), not all the variables potentially associated ARDS in AP can be evaluated and analyzed for constructing the model. Further prospective research should be done for comprehensively analyzing the association between clinical characteristics and ARDS in AP. Researches should also focus on the management including some drugs or related interventions (such as glucocorticoids, early ventilation support, blood purification, etc.) in patients with early-stage of ARDS in AP.

## 5 Conclusion

In conclusion, an intuitive nomogram with easily available laboratory parameters for the prediction of in-hospital incidence of ARDS in patients with AP was performed. The incidence of ARDS for an individual patient can be fast and conveniently evaluated by the nomogram.

## Data Availability

Datasets used and/or analyzed in the present study were availed by the corresponding author on reasonable request.

## List of abbreviations

ARDS: acute respiratory distress syndrome
AP: acute pancreatitis
SAP: severe acute pancreatitis
ICU: Intensive care unit
LOS: length of stay
SBP: systolic blood pressure
DBP: diastolic blood pressure
HR: heart rate
RR: respiratory rate
SaO2: oxyhemoglobin saturation
WBC: white blood cell count
N: neutrophil
L: lymphocyte
HCT: hematocrit
PLT: platelet
FBS: fasting blood sugar
PT: prothrombin time
APTT: activated partial thromboplastin time
ALB: albumin
ALT: alanine aminotransferase
BUN: blood urea nitrogen
SCR: serum creatinine
AMS: amylase
LPS: lipase
LDH: lactate dehydrogenase
HDL: high density lipoprotein
LDL: low density lipoprotein
TG: triglyceride
SOFA: sequential organ failure assessment
BISAP: bedside index of severity in acute pancreatitis
SIRS: systemic inflammatory response syndrome,
AUC: area under receiver operating characteristic curve
OR: odds ratio
AKI: acute kidney injury
FFA: free fatty acid

## Acknowledgments

No

## Declarations

### Ethics approval and consent to participate

The study was approved by institutional review board of Changsha Central Hospital of University of South China and the Second Xiangya Hospital of Central South University. Due to retrospective characteristics of the study, informed consent was waived.

### Author Contributions

The manuscript writing and patient’s data recording were done by Ning Ding.

Cuirong Guo, Kun Song, Changluo Li, Yang Zhou and Guifang Yang assisted in information collection. Xiangping Chai analyzed and interpreted the patients’ general indices. The final manuscript was read and ratified by all authors.

### Consent for publication

Not applicable

## References

1. Tenner S, Baillie J, DeWitt J, Vege SS: American College of Gastroenterology Guidelines: Management of Acute Pancreatitis (vol 108, pg 1400, 2013). American Journal of Gastroenterology 2014, 109(2):302–302.

2. Banks PA, Bollen TL, Dervenis C, Gooszen HG, Johnson CD, Sarr MG, Tsiotos GG, Vege SS, Acute Pancreatitis C: Classification of acute pancreatitis-2012: revision of the Atlanta classification and definitions by international consensus. Gut 2013, 62(1):102–111.

3. Abe T, Madotto F, Pham T, Nagata I, Uchida M, Tamiya N, Kurahashi K, Bellani G, Laffey JG, Investigators L-S et al: Epidemiology and patterns of tracheostomy practice in patients with acute respiratory distress syndrome in ICUs across 50 countries. Critical Care 2018, 22.

4. Fei Y, Gao K, Li W-q: Prediction and evaluation of the severity of acute respiratory distress syndrome following severe acute pancreatitis using an artificial neural network algorithm model. Hpb 2019, 21(7):891–897.

5. Dombernowsky T, Kristensen MO, Rysgaard S, Gluud LL, Novovic S: Risk factors for and impact of respiratory failure on mortality in the early phase of acute pancreatitis. Pancreatology 2016, 16(5):756–760.

6. Reilly JP, Christie JD: Is It Possible to Prevent ARDS? Jama-Journal of the American Medical Association 2016, 315(22):2403–2405.

7. Berardi G, Morise Z, Sposito C, Igarashi K, Panetta V, Simonelli I, Kim S, Goh BKP, Kubo S, Tanaka S et al: Development of a nomogram to predict outcome after liver resection for hepatocellular carcinoma in Child-Pugh B cirrhosis. Journal of Hepatology 2020, 72(1):75–84.

8. Gandaglia G, Ploussard G, Valerio M, Mattei A, Fiori C, Fossati N, Stabile A, Beauval JB, Malavaud B, Roumiguie M et al: A Novel Nomogram to Identify Candidates for Extended Pelvic Lymph Node Dissection Among Patients with Clinically Localized Prostate Cancer Diagnosed with Magnetic Resonance Imaging-targeted and Systematic Biopsies. European Urology 2019, 75(3):506–514.

9. Raith EP, Udy AA, Bailey M, McGloughlin S, MacIsaac C, Bellomo R, Pilcher DV, Anzics, Core: Prognostic Accuracy of the SOFA Score, SIRS Criteria, and qSOFA Score for In-Hospital Mortality Among Adults With Suspected Infection Admitted to the Intensive Care Unit. Jama-Journal of the American Medical Association 2017, 317(3):290–300.

10. Ranieri VM, Rubenfeld GD, Thompson BT, Ferguson ND, Caldwell E, Fan E, Camporota L, Slutsky AS, Force ADT: Acute Respiratory Distress Syndrome The Berlin Definition. Jama-Journal of the American Medical Association 2012, 307(23):2526–2533.

11. Zhang ZH: Model building strategy for logistic regression: purposeful selection. Annals of Translational Medicine 2016, 4(6).

12. Wirtz TH, Puengel T, Buendgens L, Luedde T, Trautwein C, Koch A: Diagnosis and Therapy of severe acute Pancreatitis in Intensive Care Medicine. Deutsche Medizinische Wochenschrift 2020, 145(12):850–860.

13. Fan E, Brodie D, Slutsky AS: Acute Respiratory Distress Syndrome Advances in Diagnosis and Treatment. Jama-Journal of the American Medical Association 2018, 319(7):698–710.

14. Liu S, Liu XQ, Chen SY, Xiao YX, Zhuang WD: Neutrophil-lymphocyte ratio predicts the outcome of intracerebral hemorrhage A meta-analysis. Medicine 2019, 98(26).

15. Sun YY, Chen CE, Zhang XX, Weng XC, Sheng AQ, Zhu YK, Chen SJ, Zheng XX, Lu CS: High Neutrophil-to-Lymphocyte Ratio Is an Early Predictor of Bronchopulmonary Dysplasia. Frontiers in Pediatrics 2019, 7.

16. Hayama T, Hashiguchi Y, Okada Y, Ono K, Nemoto K, Shimada R, Ozawa T, Toyoda T, Tsuchiya T, Iinuma H et al: Significance of the 7th postoperative day neutrophil-to-lymphocyte ratio in colorectal cancer. International Journal of Colorectal Disease 2020, 35(1):119–124.

17. Huang L, Chen CY, Yang LJ, Wan R, Hu GY: Neutrophil-to-lymphocyte ratio can specifically predict the severity of hypertriglyceridemia-induced acute pancreatitis compared with white blood cell. Journal of Clinical Laboratory Analysis 2019, 33(4).

18. Zemans RL, Matthay MA: What drives neutrophils to the alveoli in ARDS? Thorax 2017, 72(1):1–3.

19. Liu CN, Zhou XF, Ling LQ, Chen S, Zhou J: Prediction of mortality and organ failure based on coagulation and fibrinolysis markers in patients with acute pancreatitis A retrospective study. Medicine 2019, 98(21).

20. Akbal E, Demirci S, Kocak E, Koklu S, Basar O, Tuna Y: Alterations of platelet function and coagulation parameters during acute pancreatitis. Blood Coagulation & Fibrinolysis 2013, 24(3):243–246.

21. Gomercic C, Gelsi E, Van Gysel D, Frin AC, Ouvrier D, Tonohouan M, Antunes O, Lombardi L, De Galleani L, Vanbiervliet G et al: Assessment of D-Dimers for the Early Prediction of Complications in Acute Pancreatitis. Pancreas 2016, 45(7):980–985.

22. Papachristou GI, Muddana V, Yadav D, Whitcomb DC: Increased Serum Creatinine Is Associated With Pancreatic Necrosis in Acute Pancreatitis. American Journal of Gastroenterology 2010, 105(6):1451–1452.

23. Wan JH, Shu WQ, He WH, Zhu Y, Zhu Y, Zeng H, Liu P, Xia L, Lu NH: Serum Creatinine Level and APACHE-II Score within 24h of Admission Are Effective for Predicting Persistent Organ Failure in Acute Pancreatitis. Gastroenterology Research and Practice 2019.

24. Lipinski M, Rydzewski A, Rydzewska G: Early changes in serum creatinine level and estimated glomerular filtration rate predict pancreatic necrosis and mortality in acute pancreatitis Creatinine and eGFR in acute pancreatitis. Pancreatology 2013, 13(3):207–211.

25. Seeley EJ: Updates in the Management of Acute Lung Injury: A Focus on the Overlap Between AKI and ARDS. Advances in Chronic Kidney Disease 2013, 20(1):14–20.

26. Holder AL, Gupta N, Lulaj E, Furgiuele M, Hidalgo I, Jones MP, Jolly T, Gennis P, Birnbaum A: Predictors of early progression to severe sepsis or shock among emergency department patients with nonsevere sepsis. International Journal of Emergency Medicine 2016, 9.

27. Wiedermann CJ: Hypoalbuminemia and the Risk of Acute Kidney Injury in Sepsis. Critical Care Medicine 2019, 47(4):E377–E378.

28. Hong WD, Lin SH, Zippi M, Geng WJ, Stock S, Basharat Z, Cheng BC, Pan JY, Zhou MT: Serum Albumin Is Independently Associated with Persistent Organ Failure in Acute Pancreatitis. Canadian Journal of Gastroenterology and Hepatology 2017, 2017.

29. Yue W, Liu Y, Ding W, Jiang W, Huang J, Zhang J, Liu J: The predictive value of the prealbumin-to-fibrinogen ratio in patients with acute pancreatitis. International Journal of Clinical Practice 2015, 69(10):1121–1128.

30. Li SK, Zhang YS, Li MJ, Xie C, Wu HS: Serum albumin, a good indicator of persistent organ failure in acute pancreatitis (vol 17, 59, 2017). Bmc Gastroenterology 2017, 17.

31. Uhlig C, Silva PL, Deckert S, Schmitt J, de Abreu MG: Albumin versus crystalloid solutions in patients with the acute respiratory distress syndrome: a systematic review and meta-analysis. Critical Care 2014, 18(1).

32. Carr RA, Rejowski BJ, Cote GA, Pitt HA, Zyromski NJ: Systematic review of hypertriglyceridemia-induced acute pancreatitis: A more virulent etiology? Pancreatology 2016, 16(4):469–476.

33. Nordestgaard BG: Triglyceride-Rich Lipoproteins and Atherosclerotic Cardiovascular Disease New Insights From Epidemiology, Genetics, and Biology. Circulation Research 2016, 118(4):547–563.

34. Charlesworth A, Steger A, Crook MA: Acute pancreatitis associated with severe hypertriglyceridaemia; A retrospective cohort study. International Journal of Surgery 2015, 23:23-27.

35. Leppaniemi A, Tolonen M, Tarasconi A, Segovia-Lohse H, Gamberini E, Kirkpatrick AW, Ball CG, Parry N, Sartelli M, Wolbrink D et al: 2019 WSES guidelines for the management of severe acute pancreatitis. World Journal of Emergency Surgery 2019, 14.

36. Zhu Y, Pan XL, Zeng H, He WH, Xia L, Liu P, Zhu Y, Chen YX, Lv NH: A Study on the Etiology, Severity, and Mortality of 3260 Patients With Acute Pancreatitis According to the Revised Atlanta Classification in Jiangxi, China Over an 8-Year Period. Pancreas 2017, 46(4):504–509.

37. Guo YY, Li HX, Zhang Y, He WH: Hypertriglyceridemia-induced Acute Pancreatitis: Progress on Disease Mechanisms and Treatment Modalities. Discovery Medicine 2019, 27(147):101–109.

38. Ewald N, Hardt PD, Kloer HU: Severe hypertriglyceridemia and pancreatitis: presentation and management. Current Opinion in Lipidology 2009, 20(6):497–504.

39. Valdivielso P, Ramirez-Bueno A, Ewald N: Current knowledge of hypertriglyceridemic pancreatitis. European Journal of Internal Medicine 2014, 25(8):689–694.

40. Zhou M-T, Chen C-S, Chen B-C, Zhang Q-Y, Andersson R: Acute lung injury and ARDS in acute pancreatitis: Mechanisms and potential intervention. World Journal of Gastroenterology 2010, 16(17):2094–2099.

